# A NURSING PHILOSOPHY OF CHRONIC DISEASE SERVICES-BASED PALLIATIVE CARE NURSING DURING COVID-19

**DOI:** 10.1101/2022.10.11.22280891

**Authors:** Lailatun Nimah, Moses Glorino Rumambo Pandin, Nursalam

## Abstract

**Introduction:** Palliative care is a special service particularly in the cases of chronic disease. The scope of palliative care includes populations with old age, chronic diseases, and processes of facing death. The COVID-19 pandemic has changed the health system, including palliative care.The purpose of this study is to explain nursing palliative care during the COVID-19 pandemic.

**Method:** This paper is result of a literature review. The data bases used were Scopus, Springerlink, Science direct. The search is limited to publications in 2020-2022, open acces, and English language. Then selected using Preferred Reporting Items for Systematic Reviews and Meta-Analysis (PRISMA) diagram and obtained 13 articles.

**Result:** Palliative care as a special care aims to reduce suffering and improve the quality of life of patient and family. Palliative care can be applied to acute hospital, community, nursing care, as well as residential care homes and hospices. The goal of palliative care is to prevent and reduce suffering in any form, including reducing pain through early identification, correct assessment, as well as treatment of pain and other problems. Many barries are felt by patients and families in accessing health services during the COVID-19 pandemic due to social restrictions.

**Conclusion:** Palliative care requires the involvement of various parties, including the government, health workers, and the community to overcome various barriers. During the COVID-19 pandemic in palliative care, special health policies are needed that are able to maintain the health status of patients so that the patient’s illness does not get worse.

## Introduction

Nursing is a science that continues to grow in providing service to clients, including those suffering from chronic diseases. Palliative care is a special service particularly in the cases of chronic disease. The scope of palliative care includes populations with old age, chronic diseases, and in the processes of facing death. In this case, the focus of palliative care is to improve the quality of life of patients and their families (Wilkinson *et al*., 2018). The COVID-19 pandemic since 2019 has changed the world’s health system (Crowley and Hughes, 2021).

WHO (World Health Organization) in (Cockerham, Hamby and Oates, 2017) defines chronic disease as a long-term unrecovered disease with slow development and includes both communicable and non-communicable diseases. In this case, the risk factors that can lead to chronic disease are heart disease, stroke, cancer, chronic respiratory diseases, HIV (Human Immunodeficiency Virus)/AIDS (Acquired Immunodeficiency Syndrome), diabetes, asthma, and arthritis. Health conditions with chronic diseases at the stage of life, namely at birth, growing, working, living and aging have an impact on the social system of patients and families.

Chronic illness in a person has an impact on his work and active participation in social activities in society. Individuals with chronic diseases actually require high treatment costs in treating the disease they are suffering and create a new burden for their partners and families (Cockerham, Hamby and Oates, 2017). Chronic disease is also a challenge for health care providers. Therefore, various aspects must be considered in handling these cases, including the characteristics of the patient, the health system, and the health care provide (Braillard *et al*., 2018).

COVID-19 since 2019 has had a negative impact on individuals with chronic diseases. Fear of the spread of COVID-19 is reported to be higher in individuals suffering from chronic illnesses in Turkey. In this case, they became less focused on their work, had limited space for movement, and felt isolated from the social environment. Some employees with COVID-19 were more likely to be absent from work when they had complaints of mild coughs, flu, and other minor illnesses. They tried to adjust their working time to their health condition even though it was very difficult (Tengilimoğlu *et al*., 2022). The state of the COVID-19 pandemic has exhausted health workers, because the focus of health workers has changed for the increasing number of COVID-19 patients. Health workers are allocated less to treat palliative patients with mild symptoms. Therefore, additional health workers need to be provided in order to cope with this situation (Kates, Gerolamo and Pogorzelska-Maziarz, 2021).

Telehealth and telenursing can be treatment options during COVID-19 to treat remote patients with chronic illnesses. The provision of services through telehealth and telenursing media needs a more in-depth study and evaluation of the effectiveness and accuracy of services (Disler, Glenister and Wright, 2020). Telehealth and telenursing can be used for patients with chronic illnesses who require palliative but specialized care with minimal symptoms who can be treated at home. Resources at home must be adequate for telehealth and telenursing (Allen Watts *et al*., 2021).

Palliative care is a nursing process that is applied to chronic disease case. The stages carried out in palliative care in accordance with the National Consensus Project (NCP) include several domains, namely: a) care structure and processes; b) physical; c) psychological and psychiatric; d) social; e) spiritual, religious, and existential; f) cultural; g) taking care of patient who is facing death; and h) ethical and legal (Ferrell *et al*., 2018). Public campaigns on palliative care (including follow-up care planning and end-of-life decision making) must be carried out. The campaigns can be carried out through mass media that are adapted to age, culture, and religious or spiritual perspectives. Campaigns can be carried out on health promotion programs in the short and long term nationally. In this case, the national campaigns are expected to identify and address individual, community and system-level barriers to care (Seymour and Hons, 2018).

The impact of palliative care on the pattern of use of public health services is a reduction in emergency hospitalization, length of hospitalization, hospital costs, and increased mortality at home. Related to this, previous study revealed that communities expressed good knowledge and were willing to participate in palliative care programs (Collins *et al*., 2021). Home-based palliative care provides benefits to patients and families, namely reducing medical costs for access to hospitals, such as transportation, food costs, and health access costs. This situation facilitates access to health for patients and their families (Lustbader *et al*., 2017).

## Method

This study was a systematic review using PICO framework. The data bases used were Scopus, Springerlink, Science direct. In this case, the literatures involved were searched by topic with four keywords based on Medical Subject Heading (MeSH) and combined with Boolean operators AND and OR, as well as keywords from PICO framework include Population (chronic disease), Interest/Intervention (palliative AND COVID-19), Comparison (Nursing intervention), and Outcome (health acces). The search is limited to publications in 2020-2022, open acces, and English language.

## Result

The strategy used to search for articles using the PICO framework consists of:

**Figure 1.**
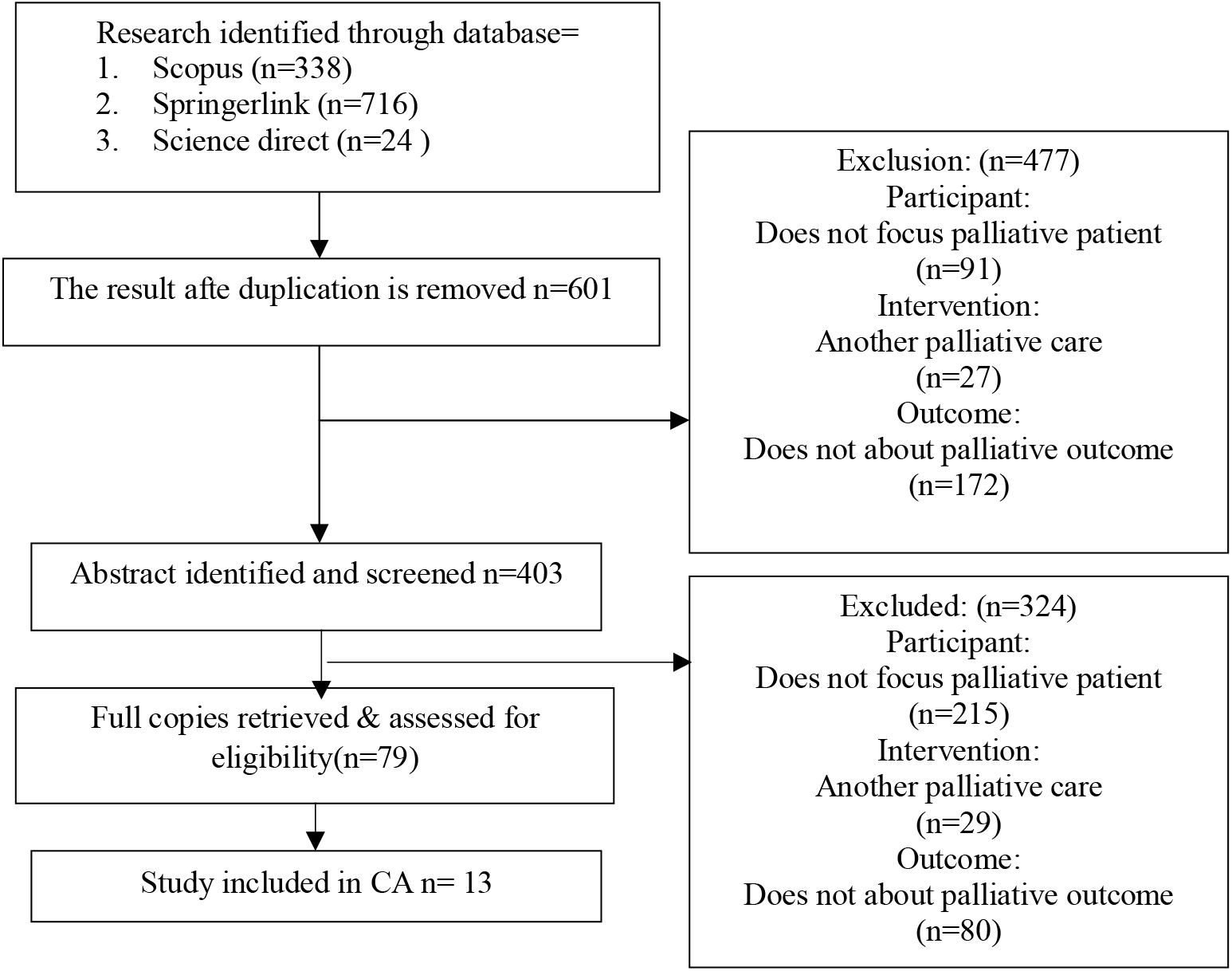
PRISMA Flow Chart

**Table 1.**
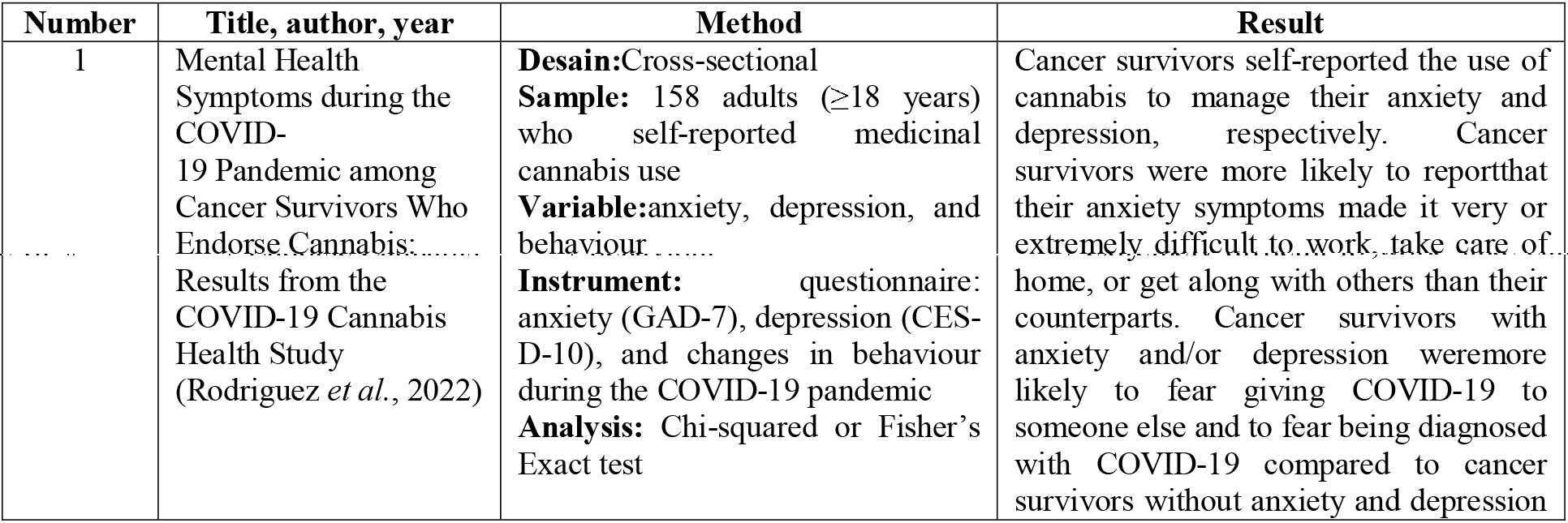
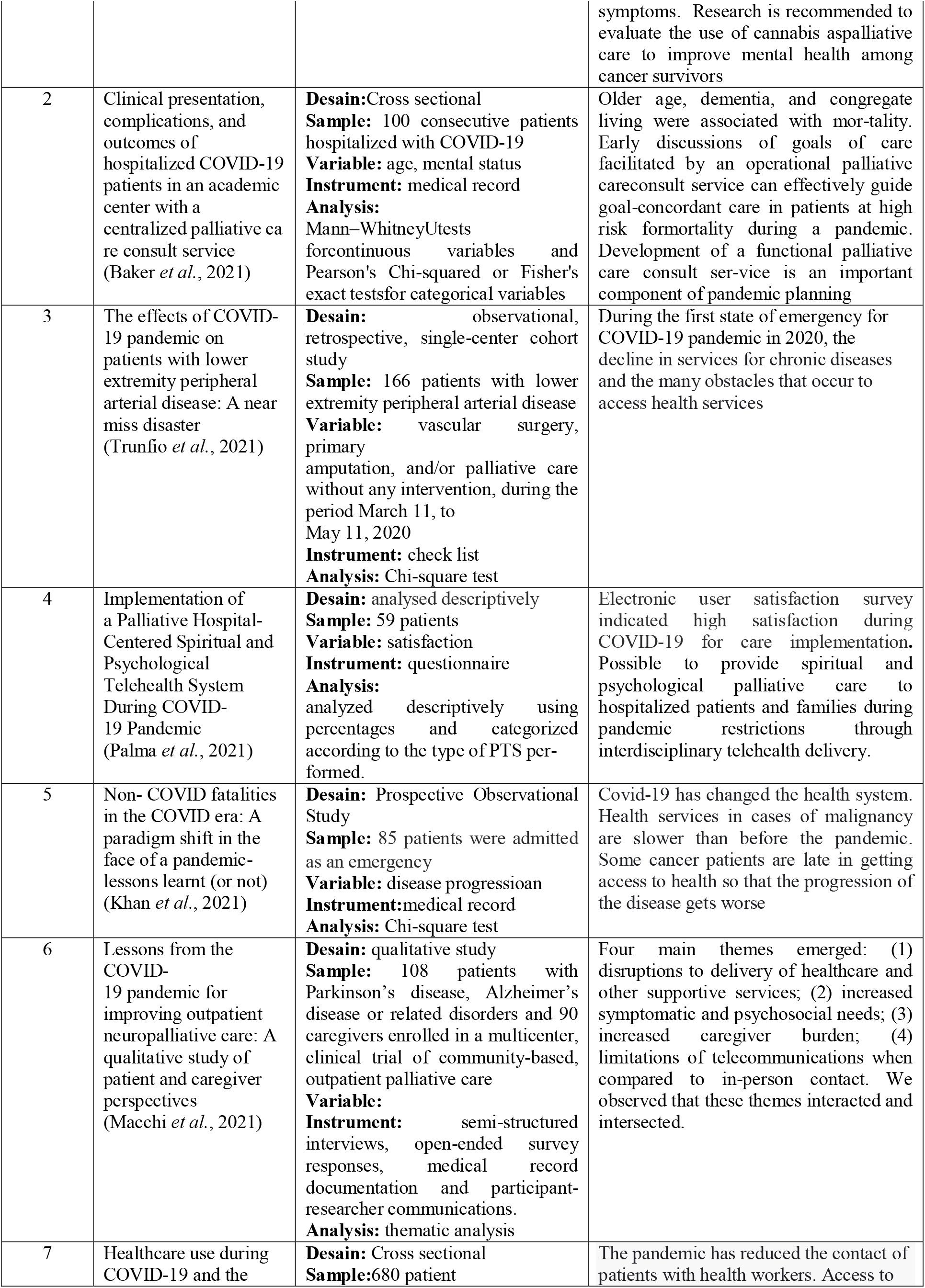
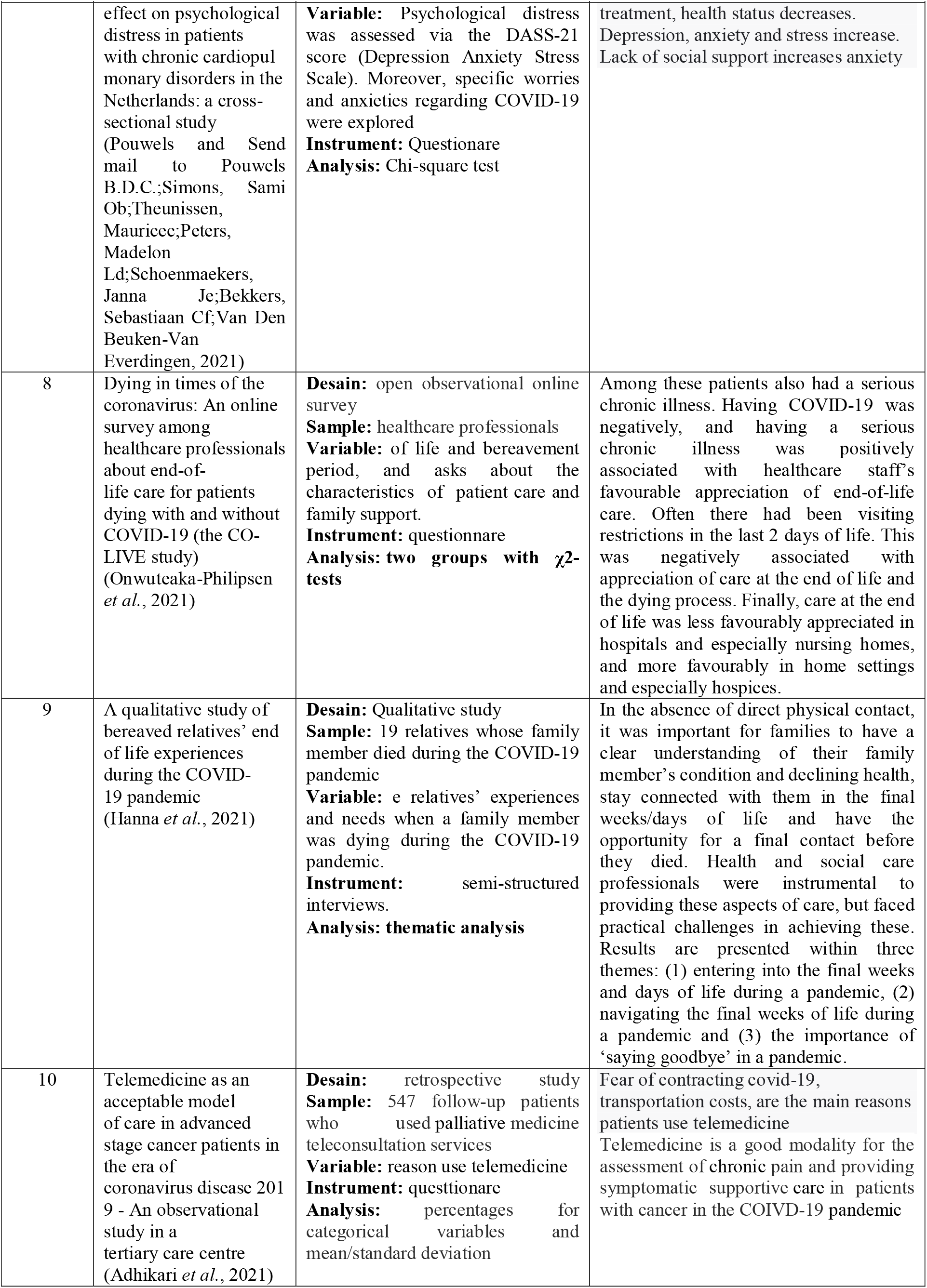
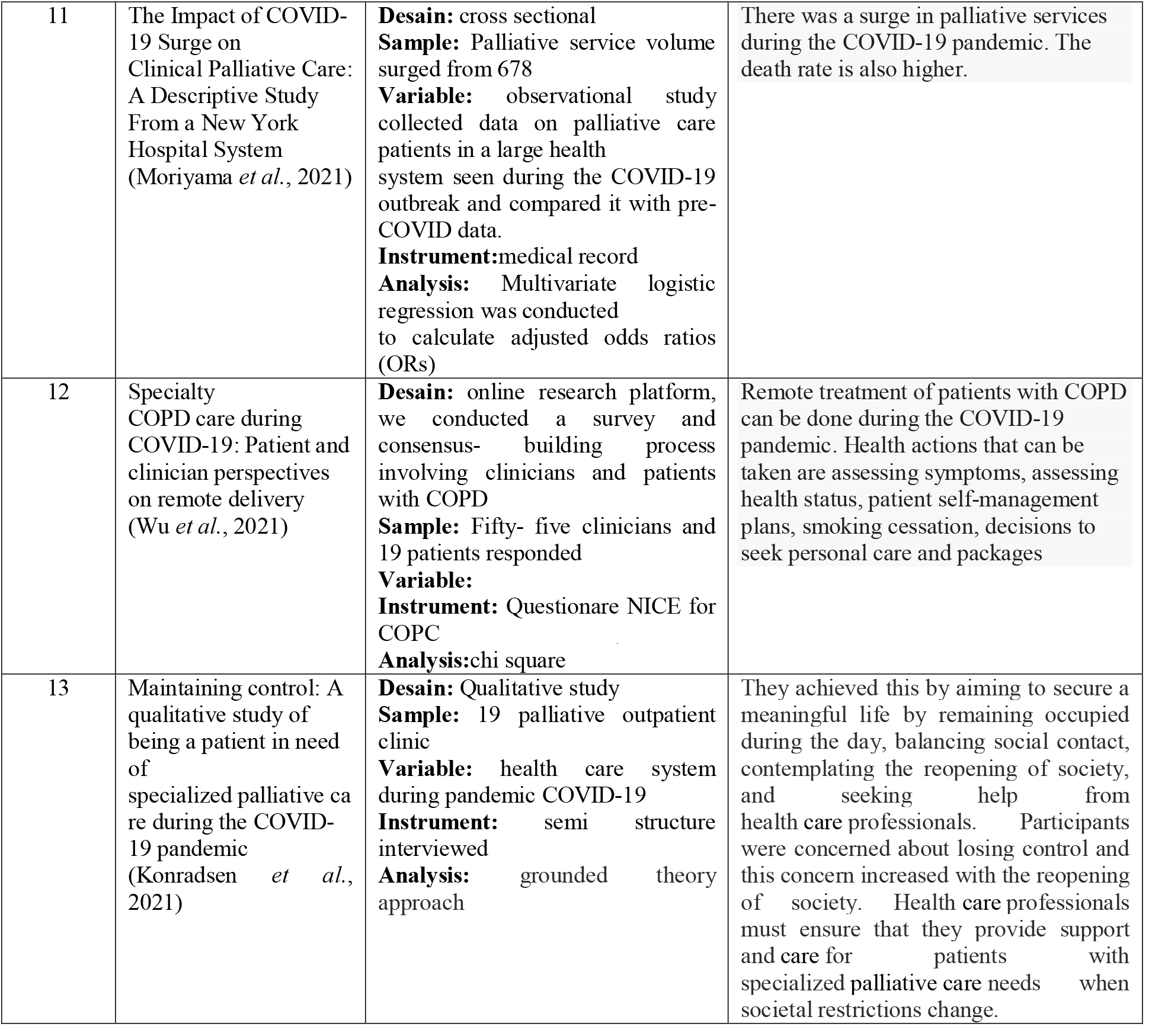
Characteristic of Reviewed Studied

## Discussion

Palliative care is treatment for adult patients, children, and families to improve the quality of life in dealing with life-threatening diseases. Scottish Partnership for Palliative Care revealed that palliative care is a basic type of care that emphasizes communication, respect for individual autonomy, and dignity. In addition, palliative care is carried out to meet the physical, social, psychological, and spiritual needs of individuals who have long-term health problems. Palliative care further also involves the role of health workers, volunteers, family, and friends.

The history of palliative care began in 1967, which was initiated by Dame Cicely Saunders who is the founder of St. Christopher in London, a hospice place used for recovering from total pain relief including the physical, emotional, social, and spiritual dimension. Elisabeth Kubler-Ross published a book in 1969 entitled On Death and Dying. The book defines five stages of grief experienced by patients with chronic illness, those are denial, anger, bargaining, depression, and acceptance. The work raised awareness about end-of-life patient care. Furthermore, Florence Wald founded America’s first hospice in 1974 after being inspired by Dr. Saunder at Yale America. The hospice movement in America is home-based and staffed by volunteers. An oncological surgeon, Dr. Balfour Mount further coined the term “palliative care” in order to distinguish it from a regular hospital in 1974.

WHO in 1990 recognized palliative care as a special care to reduce suffering and improve the quality of life of patient. A report from the Institute of Medicine published in 1997 proved that the care improved the end-of-life care with support from various philanthropic foundations. In addition, research publication in 2010 by the New England Journal of Medicine by Dr. Jennifer Temel and colleagues explained that patients with lung cancer experienced less depression and improved quality of life after receiving early palliative care. This is proven from the survival time, which was 2.7 months longer compared to patients who received standard oncology care.

Many patients’ families consider palliative care to be patient-focused and given when the patient is about to die. The level of knowledge, gender, and experience of taking care for patients with the disease greatly influence this opinion. This is supported by not all health workers receiving palliative care training in educating patients and families (Sobanski *et al*., 2020).

**Figure 2.**
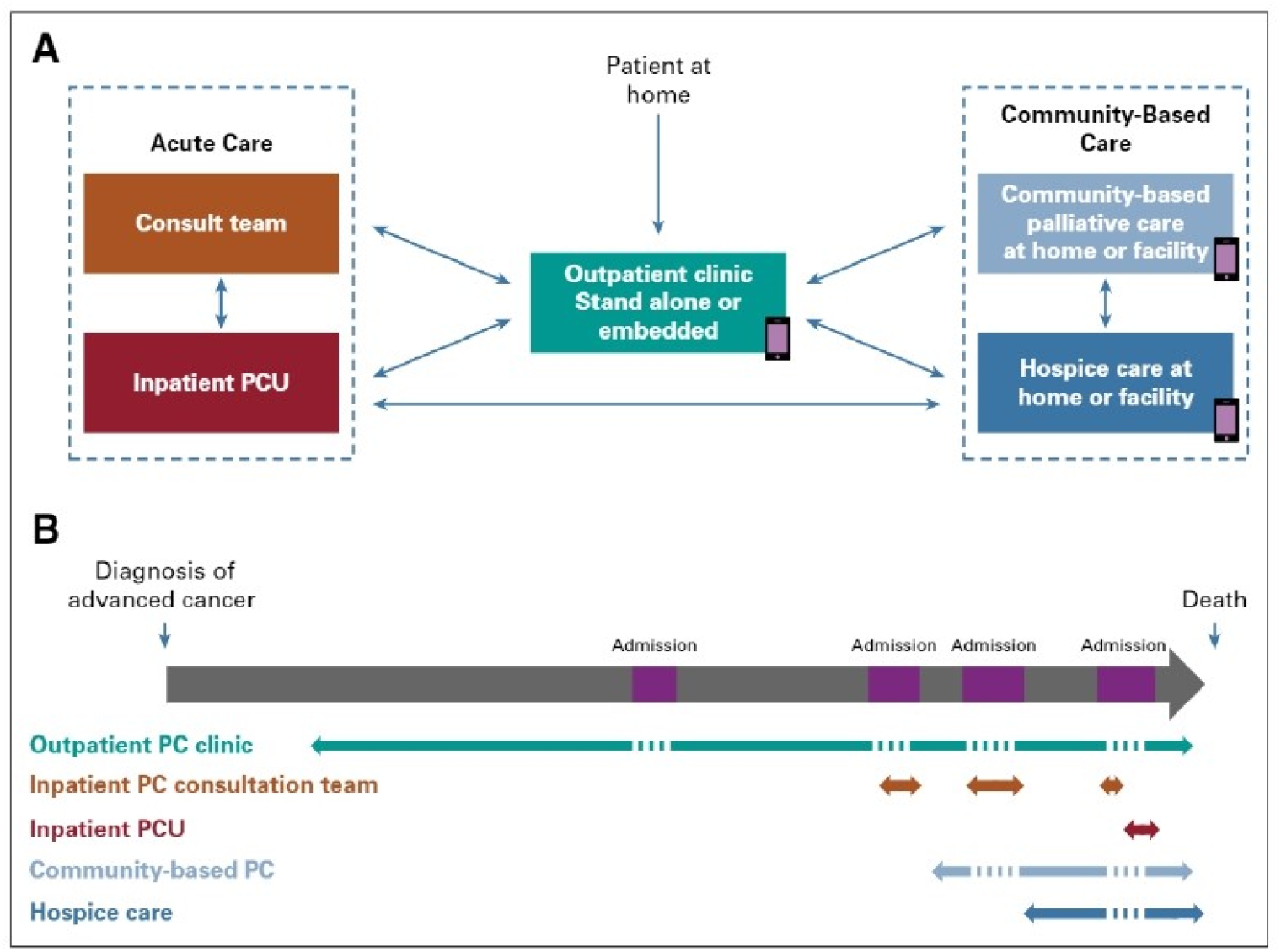
Model of specialist Palliative Care (PC) by (Hui and Bruera, 2019)

The scope of palliative care is in outpatient clinics, inpatient team consultations, acute palliative care units, community-based palliative care, and hospitals. Outpatient clinic is the main access in carrying out palliative care. Outpatient clinics can perform symptom management measures, patient health monitoring, health education, and further treatment planning. The inpatient team can act as a consultant to manage the discharge of patients who have been hospitalized. Acute palliative care can be utilized by patients with chronic disease symptoms that are difficult to manage or symptoms of more severe disease. The scope of palliative care can coordinate well to provide comprehensive services to patients and families suffering from chronic diseases (Hui and Bruera, 2019).

Outpatient palliative care clinics are palliative services for patients with the simplest chronic diseases and require fewer health resources but have a broad scope. The types of services provided include wound care, telehealth, telenursing, and home visits. Certain cases can be referred to a larger health care facility or hospitalized, namely severe physical symptoms, severe distress, severe emotional symptoms, and others. Inpatient consulting services are the main services in palliative care. The health team that is in the inpatient consultation service includes doctors and other psychosocial professionals who carry out daily monitoring in the inpatient room. Patients get more intensive program support than outpatients (Hui and Bruera, 2019).

APCU is a place to treat patients with chronic diseases that require intensive monitoring. The team assigned to this place is a special team that has received training in terms of physical, emotional, and spiritual. Medical procedures at the APCU are very diverse, including the provision of analgesics for severe pain, palliative sedation, and others. Community-based palliative programs provide home visits, home medical kits, telehealth support, and telenursing. The program consists of a healthcare team that has been defined in palliative care for minimal action. The support from family and residents around the patient’s house is needed to achieve the success of this program (Hui and Bruera, 2019). Home-based palliative care is a nursing service delivery system that focuses on patients and families in the patient’s own house environment. Family doctors and palliative nurses visit patients by maximizing the potential of the house and social environment of the house in taking care for patients with chronic diseases. Barriers can occur if the time that health workers have is limited (Mathews, Hannon and Zimmermann, 2021).

**Table 2.** Basic practice in palliative care (Hui and Bruera, 2019)

| Identifying, evaluating, diagnosing, treating, and applying solution measures for: | Identifying, evaluating, diagnosing, treating, and applying solution measures for: |
| --- | --- |
| **Physical care needs:** |  |
| a. Pain (all types) | a. Fatigue |
| b. Respiratory problems | b. Anorexia |
| c. Gastrointestinal problems | c. Anaemia |
| d. Delirium | d. Drowsiness or sedation |
| e. Wounds, ulcers, skin rash and skin lesions | e. Sweating |
| f. Insomnia |  |
| **Psychological/emotional/spiritual care needs** |  |
| a. Psychological distress | a. Spiritual needs and existential distress |
| b. Anxiety | b. Depression |
| c. Suffering of family or caregivers | c. Bereavement support for family/ caregivers |
| **Consider and manage** |  |
| **Care planning and coordination** |  |
| a. Identifying support and resources available, developing, and implementing care plan based on patients' needs |  |
| b. Providing care in the last weeks/days of life |  |
| c. Facilitating the availability and access to medications (especially opioids) |  |
| d. Identifying the psychosocial/spiritual needs of professionals providing care (including self) |  |
| **Communication issues** |  |
| a. Communicating with patient, family, and caregivers about diagnosis, prognosis, treatment, symptoms, and their management and issues related to care in the last days/weeks of life |  |
| b. Identifying and setting priorities with patient and family/caregivers |  |
| c. Providing information and guidance to patients and caregivers according to available resources |  |

Palliative care requires good interdisciplinary cooperation. Patients who suffer from chronic disease with weakness require more attention than patients with mild complaints. In this case, weakness that occurs in patients has an impact on physical and emotional stress. Patients and families often feel hopeless and apathetic about the programs provided by health workers, especially the palliative team. The quality of life for patients and families is a major improvement for the palliative team at work. Symptoms are symptoms that are often experienced by patients with chronic diseases. Giving analgesics wisely can reduce pain in patients so that the quality of life increases (Crooms and P.Gelfman, 2020). Increased knowledge and understanding of families about palliative care needs to be improved. Health education to patients involving families about what the patient wants to achieve before dying needs to be done. Good relations between health workers, patients, families must be close to set common goals (Sobanski *et al*., 2020).

The goal of palliative care is to prevent and reduce suffering in any form such as reducing pain by means of early identification, correct assessment, treatment of pain and other problems. In general, the goals of palliative are: 1) reducing pain and other distressing symptoms; 2) confirming life and death as normal processes; 3) not hastening or delaying death; 4) integrating psychological and spiritual; 4) offering a support system so that patients are as active as possible until death; 5) offering families a support system and coping with the patient’s illness and their own grief; 6) providing a team approach e.g. counseling, bereavement; 7) improving the quality of life and positively accept the disease process, 8) applying early in the course of the disease, to obtain other therapies to prolong life (Metaxa *et al*., 2021).

Palliative care is to reduce the complaints felt by patients and families in reducing the perceived adverse symptoms. Various adverse risks are decreased, can improve the quality of life and patients are able to interpret the day they spend with their family and the surrounding environment. Health costs are also the main goal that is often disputed by patients and families (Adams, 2020). Patients with cases of heart disease require shortness of breath in palliative treatment. Psychological, social, spiritual needs are also indicators of the success of palliative care in cases of heart disease. Patients always want to be near a loved one, partner, or family before the end of life. Comfort in socializing and paying attention to the patient’s dignity is the main goal in cases of heart disease (Sobanski *et al*., 2020).

The COVID-19 pandemic has an impact on the psychology of patients with chronic diseases. Many cancer patients during the COVID-19 pandemic reported symptoms of anxiety and depression and used antidepressant drugs to treat these symptoms (Rodriguez *et al*., 2022).

The contact of chronic disease sufferers with health workers is also reduced, resulting in increased stress (Pouwels and Send mail to Pouwels B.D.C.;Simons, Sami Ob;Theunissen, Mauricec;Peters, Madelon Ld;Schoenmaekers, Janna Je;Bekkers, Sebastiaan Cf;Van Den Beuken-Van Everdingen, 2021). Families caring for patients with chronic illnesses feel the burden of care has increased during the pandemic(Macchi *et al*., 2021). Social support is needed to overcome problems in patients with chronic diseases. Social restrictions during the pandemic are very detrimental to palliative care, thereby increasing depression in sufferers (Baker *et al*., 2021). Social restrictions during the period have an impact on palliative services in nursing homes due to restrictions on family visits (Onwuteaka-Philipsen *et al*., 2021).

The COVID-19 pandemic period with social restrictions requires a change in health service media, namely telehealth, telenursing and telemedicine. Patients with chronic diseases who need treatment can communicate with health workers with remote services. Fear of contracting the COVID-19 disease encourages patients and their families to use telemedicine, and telenursing and telemedicine to access health services (Adhikari *et al*., 2021). Many palliative patients report remote health services during the COVID-19 pandemic are very useful for identification of care, health education, and even spiritual and psychological services. Health services with remote systems show high satisfaction during the pandemic (Palma *et al*., 2021).The service system in hospitals during the pandemic is slower in serving palliative patients (Khan *et al*., 2021). The pandemic period has changed the health system including palliative care.

The process of death that gets medical assistance has a tremendous impact. Effective communication can reduce problems and barriers experienced by palliative health workers, patients and their families. Health workers need to communicate to patients and families regarding the medical assistance that will be provided and whether the patient and family accept or refuse. The law on the protection of medical personnel related to palliative care needs to be socialized (Mathews *et al*., 2021). The pandemic period has changed the circumstances in the process of death. Families who have family members who died due to COVID-19 cannot say goodbye and feel very lost because they cannot say goodbye due to social restrictions (Hanna *et al*., 2021).

## Conclussions

Palliative care is one of the nursing processes for patients with life-threatening illnesses that involves health workers, individuals, family, and friends. Palliative care has dimensions of physical care needs, psychological/emotional/spiritual care needs, care planning, as well as coordination and communication issues. The goal of palliative care is to prevent and reduce suffering by early identification, proper assessment, treatment of pain, and other problems. In this case, palliative care requires the involvement of various parties, namely the government, health workers and the community to overcome various barriers. During the COVID-19 pandemic in palliative care, special health policies are needed that are able to maintain the health status of patients so that the patient’s illness does not get worse.

## Data Availability

All data produced in the present study are available upon reasonable request to the authors
All data produced in the present work are contained in the manuscript
All data produced are available online at

## Acknowlegments

Thanks to all of my friends who have supported the completion of this article

